# Attrition from HIV treatment after enrollment in a differentiated service delivery model: a cohort analysis of routine care in Zambia

**DOI:** 10.1101/2022.05.30.22275759

**Authors:** Youngji Jo, Lise Jamieson, Bevis Phiri, Anna Grimsrud, Muya Mwansa, Hilda Shakwelele, Prudence Haimbe, Mpande Mukumbwa-Mwenechanya, Priscilla Lumano Mulenga, Brooke E Nichols, Sydney Rosen

**Author notes:** Corresponding author: Sydney Rosen, Boston University School of Public Health, 801 Massachusetts Ave, Boston, MA, 02118, USA. These authors have contributed equally to the work (Brooke E Nichols, Sydney Rosen).

## Abstract

**Background:** Many sub-Saharan Africa countries are scaling up differentiated service delivery (DSD) models for HIV treatment to increase access and remove barriers to care. We assessed factors associated with attrition after DSD model enrollment in Zambia, focusing on patient-level characteristics.

**Methods:** We conducted a retrospective record review using electronic medical records (EMR) of adults (≥15 years) initiated on antiretroviral (ART) between 01 January 2018 and 30 November 2021. Attrition was defined as lost to follow-up (LTFU) or died by November 30, 2021. We categorized DSD models into eight groups: fast-track, adherence groups, community pick-up points, home ART delivery, extended facility hours, facility multi-month dispensing (MMD, 4–6-month ART dispensing), frequent refill care (facility 1-2 month dispensing), and conventional care (facility 3 month dispensing, reference group). We used Fine and Gray competing risk regression to assess patient-level factors associated with attrition, stratified by sex and rural/urban setting.

**Results:** Of 547,281 eligible patients, 68% (n=372,409) enrolled in DSD models, most commonly facility MMD (n=306,430, 82%), frequent refill care (n=47,142, 13%), and fast track (n=14,433, 4%), with <2% enrolled in the other DSD groups. Retention was higher in nearly all DSD models for all dispensing intervals, compared to the reference group, except fast track for the ≤2 month dispensing group. Retention benefits were greatest for patients in the extended clinic hours group and least for fast track dispensing.

**Conclusion:** Although retention in HIV treatment differed by DSD type, dispensing interval, and patient characteristics, nearly all DSD models out-performed conventional care. Understanding the factors that influence the retention of patients in DSD models could provide an important step towards improving DSD implementation.

## Introduction

Although access to antiretroviral treatment (ART) for HIV is now widespread, ART programs worldwide continue to face the challenge of retaining patients in lifelong care. Studies in sub-Saharan Africa (SSA) suggest that only 67% of patients remain in ART programs after five years, with loss to follow-up (patients with unknown outcomes) accounting for 33% of all attrition(1).

One solution to this high attrition from ART programs has been the introduction of differentiated service delivery (DSD) models. DSD models aim to improve long-term ART retention by removing barriers to care, making service delivery more patient-centered(2), generate greater patient satisfaction, reduce costs to patients (and to providers in some cases), and create efficient and convenient service delivery(3). DSD models differ from conventional HIV care in the location of service delivery, frequency of interactions with the healthcare system, cadre of provider involved, and/or types of services provided(4). The attractiveness of DSD models is generally considered to be conditional on maintaining at least equivalent clinical outcomes to conventional care, but there remains relatively little evidence on ART retention among patients enrolled in DSD models as part of large-scale routine care in SSA(5, 6).

Zambia, a high-HIV burden country with more than 1.5 million people living with HIV and more than 81% of those individuals on ART, has rapidly scaled up a variety of DSD models(7). Participation in DSD models and ART treatment outcomes are documented in Zambia’s national electronic medical record (EMR) system, known as SmartCare. We used the SmartCare data set, the largest dataset of its kind available on DSD model uptake and outcomes, to compare patient outcomes in DSD models to conventional care and assess patient-level factors associated with retention after enrolment in different DSD models.

## Methods

### Study population and setting

The Zambian Ministry of Health (MOH) authorized the non-governmental organizations supporting HIV treatment scale up in the country to pilot various models of service delivery starting in 2014(8). Nationwide scale-up DSD models was underway by 2017. Most public sector healthcare facilities now provide at least one alternative to conventional care, with many offering multiple options. Some DSD models are described in national ART guidelines and can potentially be offered by all facilities(9)(10). Others were designed and introduced by non-governmental partner organizations working with specific facilities. Under current guidelines, to be eligible for DSD enrollment patients must be “stable” or “established on treatment,” defined as having been on first-line ART for at least 6 or 12 months and having demonstrated viral suppression.

The SmartCare database covers approximately three-quarters of all ART patients in Zambia, with the remainder accessing care at clinics that do not utilize the system(11). Routine medical record data are either entered into SmartCare portals in real-time during the patient interaction or transcribed from paper records, typically within a day or two of the interaction. With the assistance of the national MOH, we accessed a subset of data fields from the entire SmartCare cohort from the study period from January 1, 2018 to November 30, 2021.

We defined a cohort of all patients, aged 15 years or older, who initiated ART on or after January 1, 2018 and had been retained in care ≥6 months after ART initiation. We then categorized those patients into 14 different models of care based on data recorded in SmartCare, including the number of months of ART dispensed. These DSD models were grouped into eight relatively homogeneous analytic groups: facility fast track services, adherence groups, community pick-up points, home ART delivery, extended clinic hours, and models based on dispensing duration alone (Table 1). Enrolment of patients into specific models depended on both national eligibility guidelines and facility characteristics. In general, facilities assigned individuals to a model of care based on model availability, patient characteristics and preferences, and operational considerations. Transfer to a different DSD model was permitted, but transfers may not have been captured fully in the electronic medical records during the study period and thus cannot be accounted for in our analysis.

**Table 1.** Differentiated service delivery models in Zambia and analytic groups.

| **Analytic group** | **Specific DSD models included in the group** | **Description** |
| --- | --- | --- |
| Fast track | Fast-track services at facility | A model that creates a separate queue, kiosk, or procedure at a facility to speed up service delivery for stable patients. In Zambia, this typically involves a separate and shorter queue for quick dispensing when a full clinical visit is not indicated. |
| Adherence groups | Community adherence groups (CAG), urban adherence groups, rural adherence groups | Community adherence groups: groups of $\pm 6$ people, based on residential proximity or patient preference, meet monthly at a designated place in the community. Members collect medication at clinical appointments for other CAG members, in a rotating fashion.<br>Rural/Urban adherence groups: groups of 15-30 people receive group adherence counselling and pre-packed ART dispensation by a healthcare worker or community health workers outside of typical clinic hours. |
| Community pick-up points | Central dispensing units, community ART distribution points, community retail pharmacy, health post, mobile ART distribution | Any model that delivers ART to pick-up points outside clinic facilities, such as medication lockers, community pharmacies, central dispensing units, community ART distribution points, health posts (remote facilities that do not typically dispense ART), mobile ART distribution (van-based clinic). |
| Home ART delivery | Home ART delivery | Trained community health workers (CHWs) linked to facilities conduct home visits to deliver ART, conduct health screening, monitor adherence, and refer patients as required. All community services are captured on a tablet-based SmartCare app. |
| Extended clinic hours | After/before clinic hours, scholar model, weekend clinics | ART dispensing available outside standard clinic hours, either before or after hours on weekdays, expanded hours for school-going youth, or ART dispensing on the weekend. |

| Analytic group | Specific DSD models included in the group | Description |
| --- | --- | --- |
| Facility multi-month dispensing (MMD) | 4-6-month dispensing of ART at a facility and not in any other DSD model | Any model in which the primary goal is to dispense medications for a longer duration than is done under conventional care (4 to 6 months). Dispensing is typically done at a standard facility visit. Those categorized as “facility MMD” in this study were not reported as participating in any other DSD model. |
| Frequent refill care | 1-2 month dispensing of ART at a facility | As 3-month dispensing has expanded, patients receiving 1-2-month refills may be considered higher risk in some way, though inventory shortages could also play a role. |
| Conventional care | 3-month dispensing of ART at a facility | At the time of the study, the widely followed standard of care was to provide a 3-month supply of ART at quarterly clinic visits. 3-month dispensing was originally considered a differentiated model; we refer to it as “conventional” to emphasize that 3-month dispensing is our reference model. |

Source: Table modified from (7)

In reviewing the dataset, we observed that many patients who received 4-6 months of ART medications, known as multi-month dispensing (MMD), at their most recent clinic visit, these were not labeled as MMD in Smartcare. We therefore assigned patients to analytic groups as follows. Patients assigned to any DSD model except MMD were assigned to the relevant analytic group shown in Table 1. Patients assigned to MMD were combined with patients not assigned to any DSD model (remaining in conventional care). This combined population was then stratified by the duration of dispensing at their most recent clinic visit, with dispensing intervals defined as 1-2 months (“frequent refill care”), 3 months (conventional care), or 4-6 months (facility MMD). We refer to patients we assigned to facility MMD, frequent refill care, or conventional care as being “enrolled” in these models, even though most were not identified as such in SmartCare. We also note that 3-month dispensing (3MD) was originally regarded as a differentiated model in Zambia, to distinguish it from the earlier standard of care, which allowed only 1-2 month dispensing. Over the course of the study period, however, 3MD was widely implemented as standard care. For this analysis, we therefore refer to 3MD as “conventional care,” while 1-2-month dispensing is labeled as “frequent refill care,” and only 4-6-month dispensing is described as “multi-month” (facility MMD). All models of care could have different number of months dispensing, aside from “frequent refill care” “conventional care” and “facility MMD”, as these were defined in terms of dispensing duration and location of dispensing alone.

### Outcomes and data analysis

Our primary outcome was attrition from care at any time between January 1, 2018 and November 30, 2021. Attrition was defined as patients who were reported to be lost to follow-up (not found >28 days from last scheduled appointment), had stopped ART (patient found after ≤ 28 days but stopped medications), or had died. We note that because the data censoring date, November 30, 2021, was common to all patients, the follow-up duration for each patient in the cohort depended on the date the patient initiated ART (any time on or after January 1 2018).

Using Fine and Gray competing risk regression(12), we estimated the hazard ratios of attrition, with transfer to a different facility considered as a competing risk of the attrition event of each DSD model group compared to conventional care. We adjusted for age, sex, location (urban vs rural), and care level (health post, primary clinic, hospital). Results were stratified by most recent ART dispensing interval. We conducted a secondary analysis further stratifying results by location (urban or rural) and sex.

### Ethics review

This study was approved by the ERES Converge IRB (Zambia), the Human Research Ethics Committee (Medical) of the University of Witwatersrand (South Africa), and the Boston University Institutional Review Board (USA). The requirement for informed consent was waived for this study, which was a review of retrospective medical record data only.

## Results

### Study population and DSD model enrollment

The full data set included records for 1,278,627 individual patients receiving care at 1,486 facilities located in 93 districts and 10 provinces. Of these, 547,281 patients were eligible for analysis, as shown in Fig 1. During the study period, 68% (n=372,409) enrolled in DSD models, most commonly facility MMD (n=306,430, 82%), frequent refill care (n=47,142, 9%) or fast track (n=14,433, 4%). The remaining patients were distributed among extended clinic hours (0.4%), home ART delivery (0.3%), community pick-up points (0.3%), and adherence groups (0.2%).

**Fig 1.**
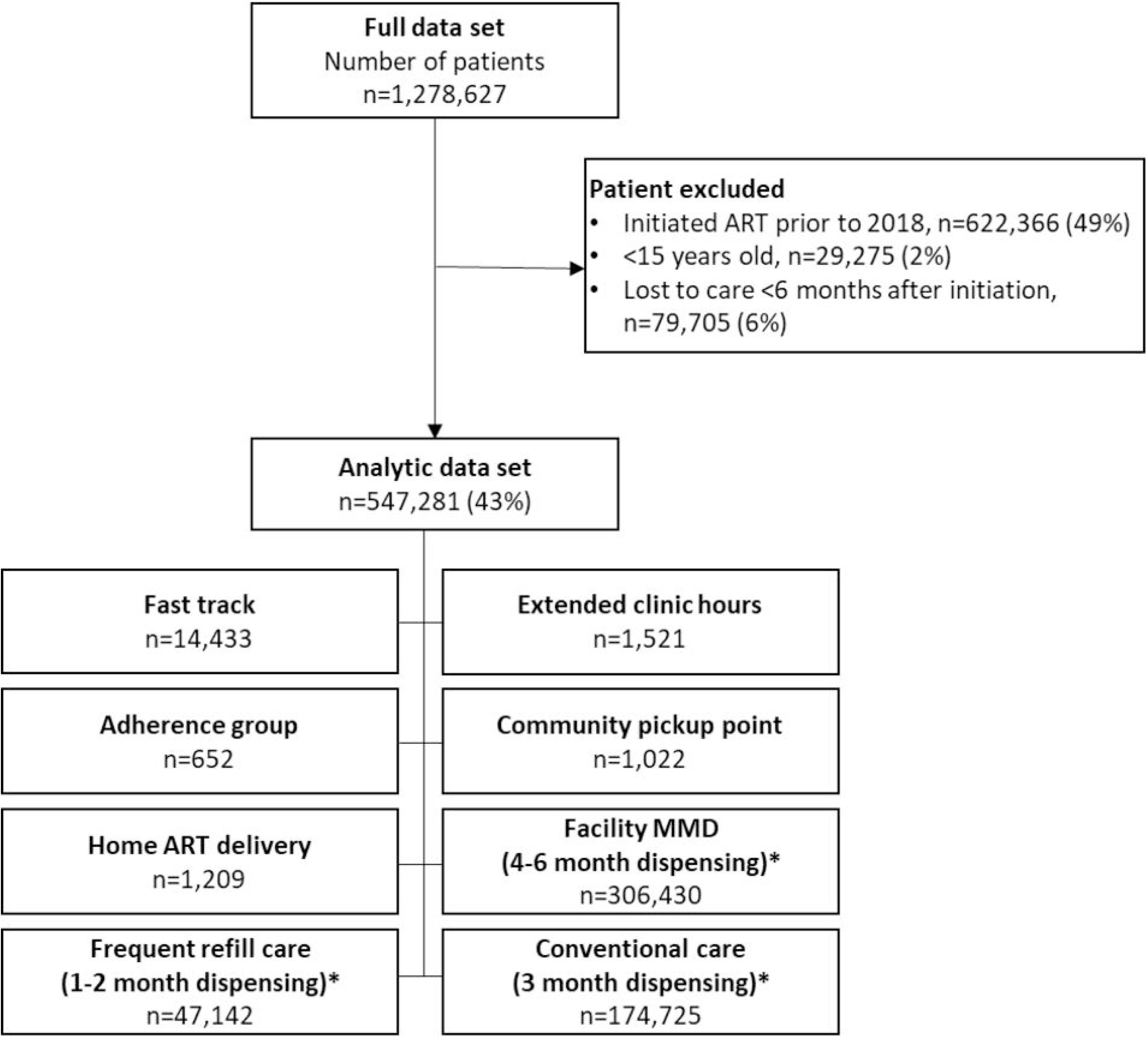
Composition of analytic data set. *Facility MMD, frequent refill care, and conventional care as defined in manuscript; all other models are indicated in SmartCare database.

Consistent with the national ART program as a whole (9), the majority of patients enrolled in DSD models were female (61%, n=228,442) (Table 2). The cohort had a median (IQR) age of 36 (29-44) years and was mostly based in urban areas (n=218,260, 59%). Age and sex distributions were similar between DSD model types, except in the case of frequent refill care and extended clinic hours, which had a larger proportion of young adults (15-24 years) than did the other models. Most study participants (59%) were dispensed between 4 and 6 months of ART at their last clinic visit, regardless of DSD model.

**Table 2.**
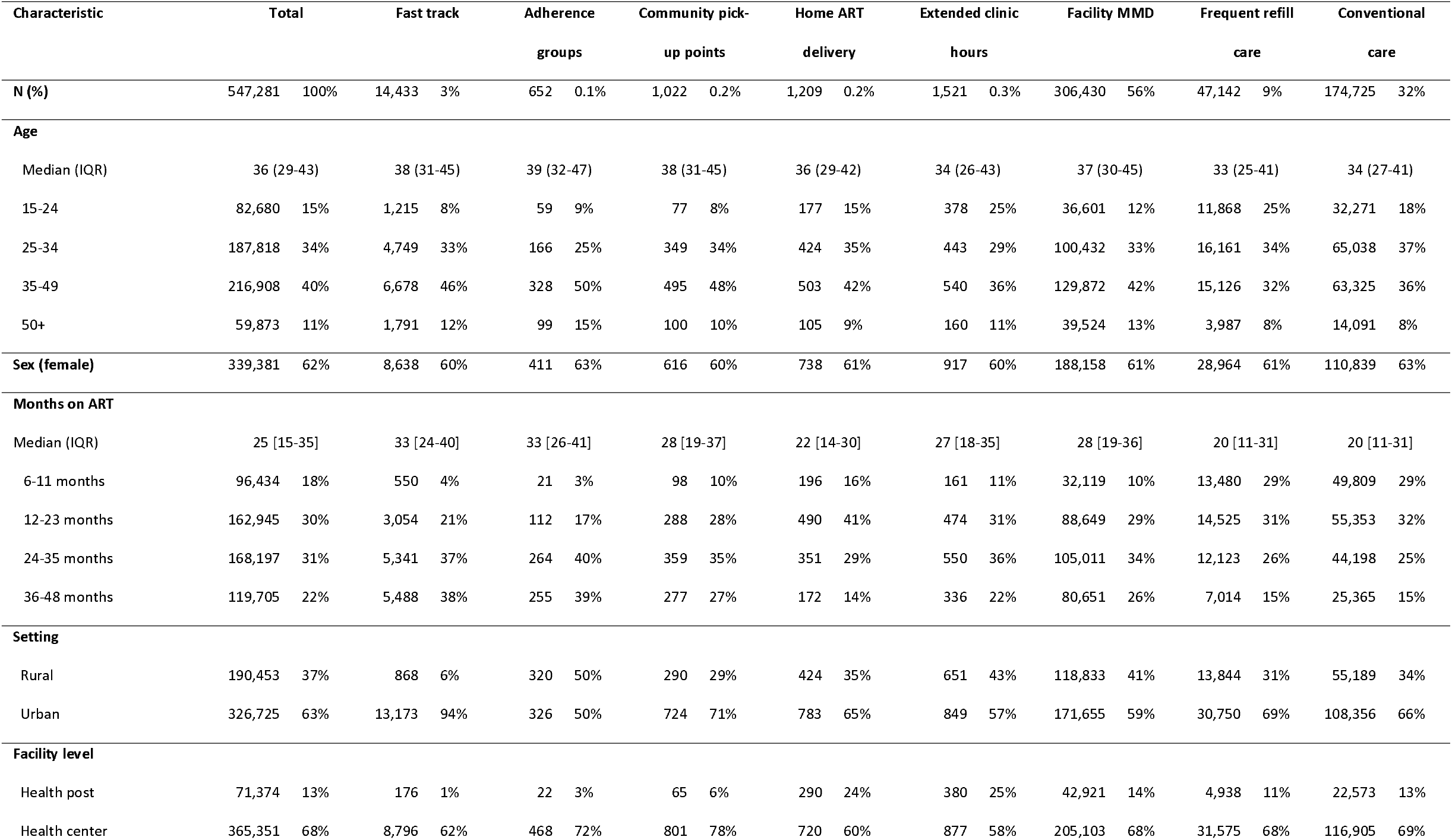
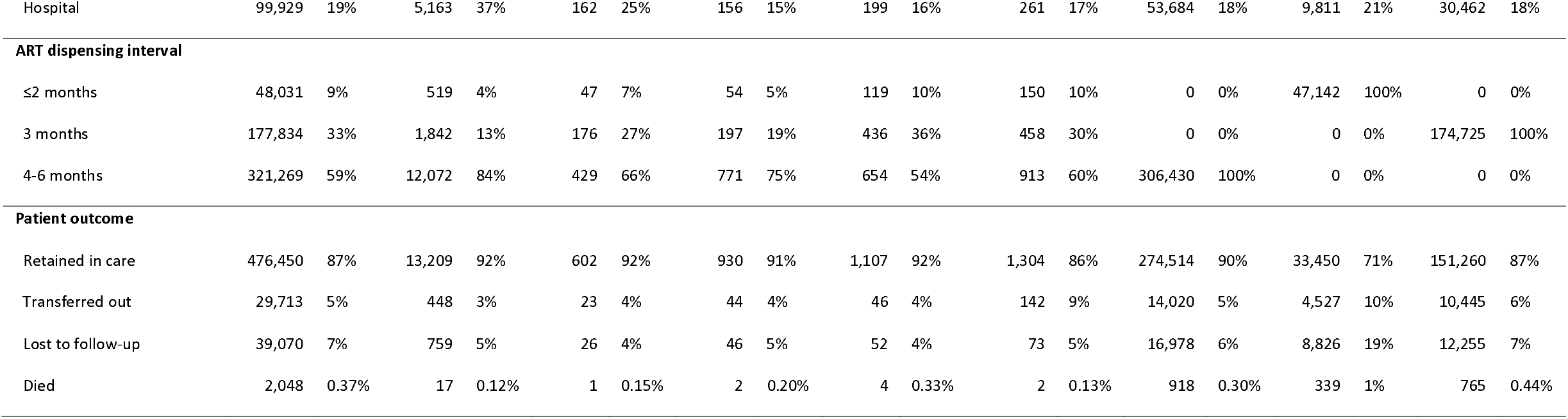
Characteristics of patients who initiated ART treatment on or after Jan 1, 2018 and were retained in care ≥ 6 months in Zambia.

### Treatment outcomes

Retention in DSD models was high (86-92%) within the study period for all models including conventional care, with the exception of frequent refill care, where retention was substantially lower at 71% (Table 2). Transferred out was higher for frequent refill care (10%) and extended clinic hours (9%) groups compared to other care models (3-6%); loss to follow-up was especially high for frequent refill care (19%) than any other care models (4-7%); <1% of patients were reported to have died in any model during the follow-up period.

Across all dispensing intervals, patients in SmartCare-designated DSD models had significantly lower risk of attrition compared to those in the respective reference model (frequent refill care for ≤ 2 months’ dispensing, conventional care for ≤ 3 months, and facility MMD for 4-6 months), with the exception of fast track models with ≤2-month dispensing, which had very slightly higher attrition (Fig 2). Patients enrolled in extended clinic hours models had a significantly lower risk of attrition than the respective reference groups, with an adjusted hazard ratio (aHR [95% confidence interval]) ranging from 0.36 [0.22-0.61] for patients receiving ≤2 months of ART to 0.71 [0.51-0.99] for patients receiving 4-6 months of ART. Patients utilizing extended clinic hours were also more likely to transfer care to other facilities than were those enrolled in other models. Attrition rates and patterns are generally similar between 3-month and 4–6-month dispensing.

**Fig 2.**
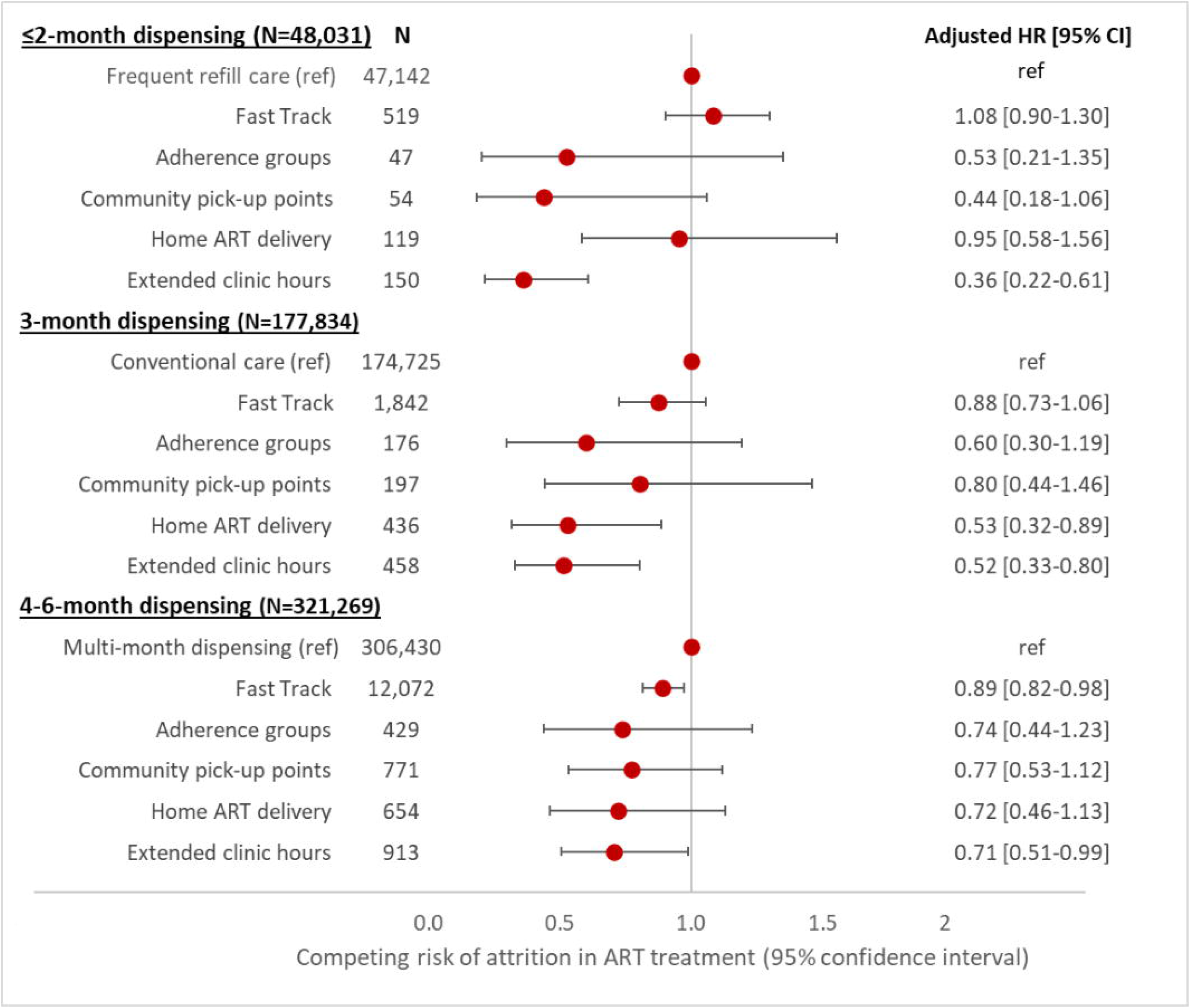
Adjusted hazard ratio for attrition from care, stratified by ART dispensing interval.

Fig 3 presents the results of our regression analysis stratified by sex and location. For the urban subpopulations, there was no statistically significant difference in risk of attrition among most DSD models except the extended clinic hours model. For the rural subpopulations, frequent refill care was associated with a statistically greater risk of attrition, with point estimates for aHR ranging between 1.70 and 1.91, while MMD was associated with a statistically lower risk of attrition in both sexes (aHR ranging between 0.77 and 0.81), compared to conventional care. For the rural female subpopulation, there was no substantial difference in risk of attrition among other DSD models compared to conventional care. For the rural male subpopulation, however, community pick-up point and extended clinic hours are associated with a significantly lower risk of attrition of 0.13 [0.02-0.93] and 0.49 [0.26-0.90], respectively, compared to conventional care.

**Fig 3.**
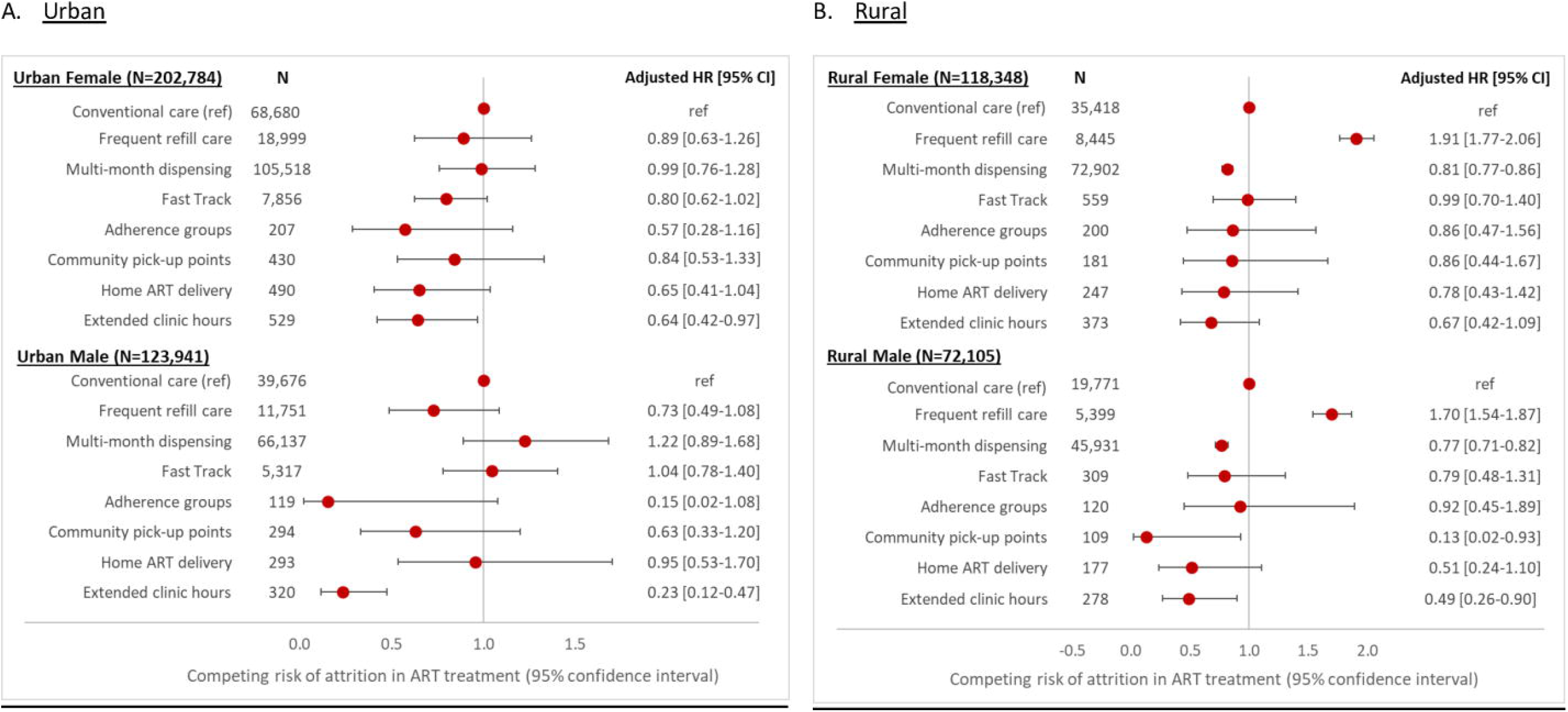
Adjusted* hazard ratio of attrition, stratified by location and gender. *Adjusted by age and facility level.

## Discussion

In this analysis, we found that the risk of attrition from care in the period from 2018-2021 in Zambia differed by service delivery model and ART dispensing interval. Importantly, retention in care was high across seven of eight models, ranging from 86% to 92%. The one exception, frequent refill care, with 71% retention, is unsurprising given the likelihood that patients deemed to be at higher risk of loss to follow-up or death are likely to be dispensed ART at shorter intervals intentionally to ensure frequent clinic follow-up for more regular monitoring. Despite a relatively lower retention rate (86%) in the extended clinic hours model, patients still maintain a statistically better retention than those remaining in conventional care across all dispensing intervals after accounting for transfer as a competing risk.

When we analyzed subpopulations by setting and sex, we found that the risk of attrition did not vary significantly by DSD model among urban patients. Rural males appear to be benefiting most from DSD enrollment. Of note is the relatively high hazard of attrition among rural patients enrolled in frequent refill care. As mentioned above, patients receiving frequent refills include those who are not regarded as stable on ART and are believed to require frequent clinical monitoring. At the same time, however, having to make repeated visits to a healthcare facility may be particularly challenging for rural patients, for whom travel distances tend to be greater. The possibility that the frequent refill model encourages attrition from care, rather than improving the quality of care, should be considered.

Our findings are generally consistent with those of other studies of DSD model outcomes in SSA(6). Previous studies found high retention of patients in adherence clubs and home-based care models in South Africa and Kenya, among other countries(13, 14). Studies in Zambia have showed that DSD models achieve comparable or better outcomes than conventional care(15, 16). In ART programs in SSA, males have traditionally been at higher risk of attrition than females(17), but we found that rural males in most DSD models in Zambia were retained in care as well as or relatively better than females in the same models and settings.

Other studies have also suggested that while DSD models may not improve retention among currently eligible patients, who are already “stable” at the time of DSD model enrollment and thus likely to continue to demonstrate high retention, it can still offer other benefits to patients and providers by reducing costs and improving quality(18)(19). If there is no significant difference in the risk of attrition between DSD models and conventional care (as shown in the urban female group in Fig 3), DSD model choice can be based on other factors such as operational feasibility or cost to the provider or patient.

Our study had several limitations. First, and most important, it was an observational study, and we know that patients were not enrolled in DSD models at random. It is likely that patients offered DSD model enrollment were believed by facility staff to be “good adherers,” while those thought to be at higher risk of attrition were held in frequent refill care or conventional care, where they could potentially be monitored more closely. Similarly, the same model may be assigned to different dispensing intervals for a slightly different population of patients(14). For example, the different retention levels observed in fast track (i.e., lower retention for ≤2 month and better retention for 3 and 4-6 month dispensing groups) reflect the patient population served rather than the model of care itself. Third, the data used was routinely-collected patient record data. We guess that recording of patients’ entry into DSD models was incomplete; it is likely that some patients in DSD models were not reported as such. The database, moreover, did not contain information on adverse events, waiting time at facilities, staff shortages, drug supply issues, or patient travel distance – all factors which could affect patient attrition. Finally, while DSD enrollment in our study occurred prior to the COVID-19 pandemic, retention over the course of 2020 may have been affected both by pandemic restrictions (limitations on travel, etc.) and by pandemic adaptations, such as more emphasis on out-of-clinic service delivery and multi-month dispensing.

## Conclusions

Despite the limitations described above, this study provides evidence that most of the differentiated service delivery models for HIV treatment in use in Zambia between 2018 and 2021 were associated with substantial and consistent improvements in retention in care. DSD models of care that are not demonstrating favorable retention rates should be reconsidered for further implementation or redesigned to meet the needs of the populations they serve. The strategic design and targeting of DSD models are critical to their success in retaining patients on ART. Understanding the factors that influence the retention of ART patients in DSD models could provide an important step towards improving DSD implementation.

## Data Availability

Raw data were obtained from Zambia's national electronic medical record system. Derived data supporting the findings of this study are available from the corresponding author [SR] on request.

## Supporting information

S1 Fig. Cumulative incidence curves by stratified groups

## Authors’ contributions

SR and BEN conceived the study. YJ, LJ, BEN designed the study. BP, MM, HS, PH, MMM, PLM led study data collection. YJ, LJ analyzed the data, and SR, BEN contributed to data analysis. YJ, LJ, SR, BEN wrote the first draft of the manuscript. All authors reviewed and edited the manuscript. All authors have read and approved the final manuscript.

## Acknowledgements

None.

