## Supplemental figures for "Attrition from HIV treatment after enrollment in a differentiated service delivery model: a cohort analysis of routine care in Zambia"

**S1 Fig. Cumulative incidence curves by stratified groups**

We used a competing risk regression analysis and the risk of attrition was calculated by accounting transferred out as a competing risk of attrition. Below are the cumulative incidence curves of attrition and transferred out over time (months) by stratified groups; most DSD models show a similar or greater proportion of attrition than the proportion of transferred out while the extended clinic hours model (and community pick-up points in some subgroup) show a greater proportion of attrition than the proportion of transferred out. These resulted in a significant reduction of relative risk of attrition compared to the reference group in the regression analysis. (Figure 2 and 3)

1) **≤2 months dispensing**

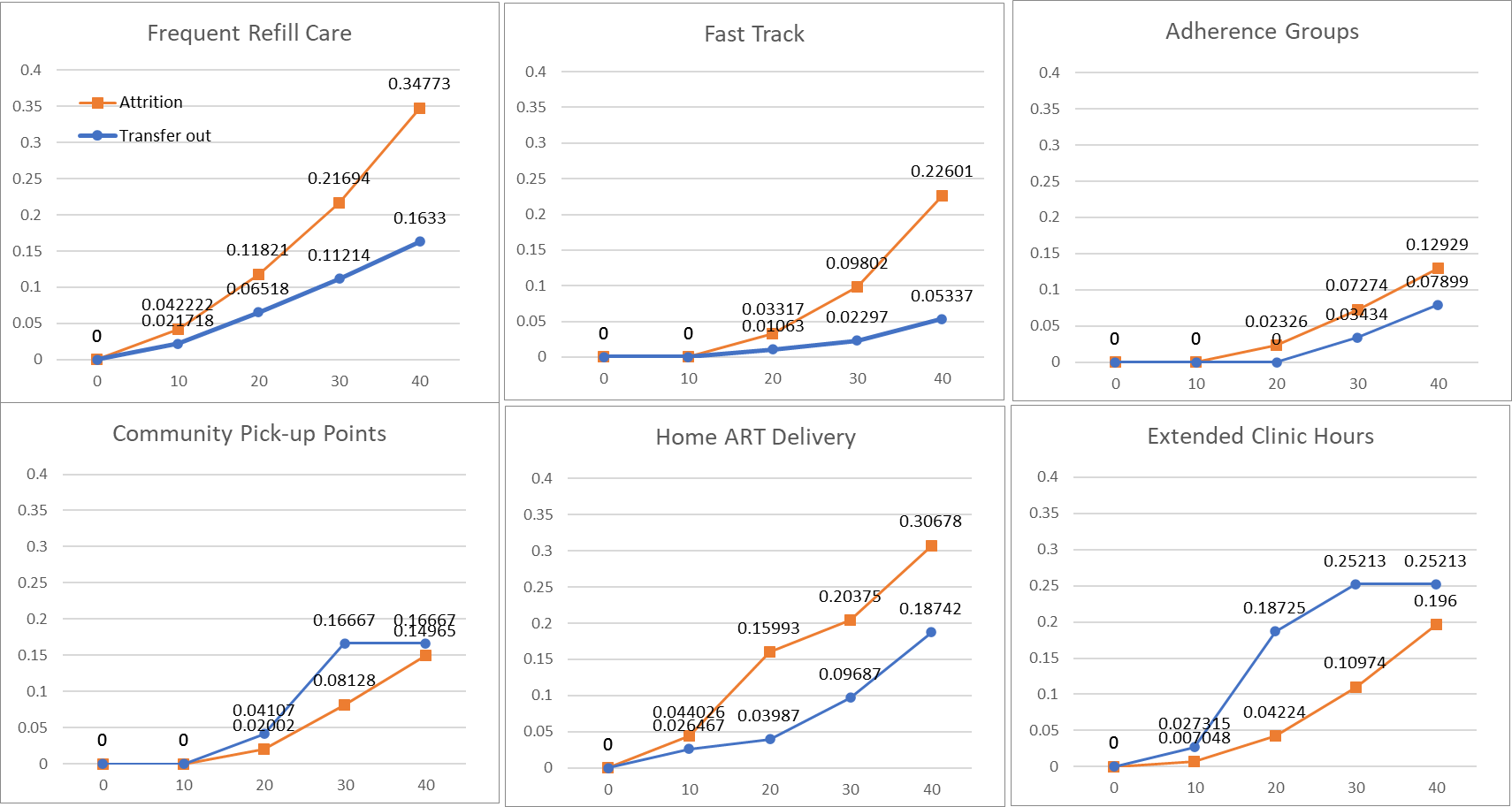

2) **3 months dispensing**

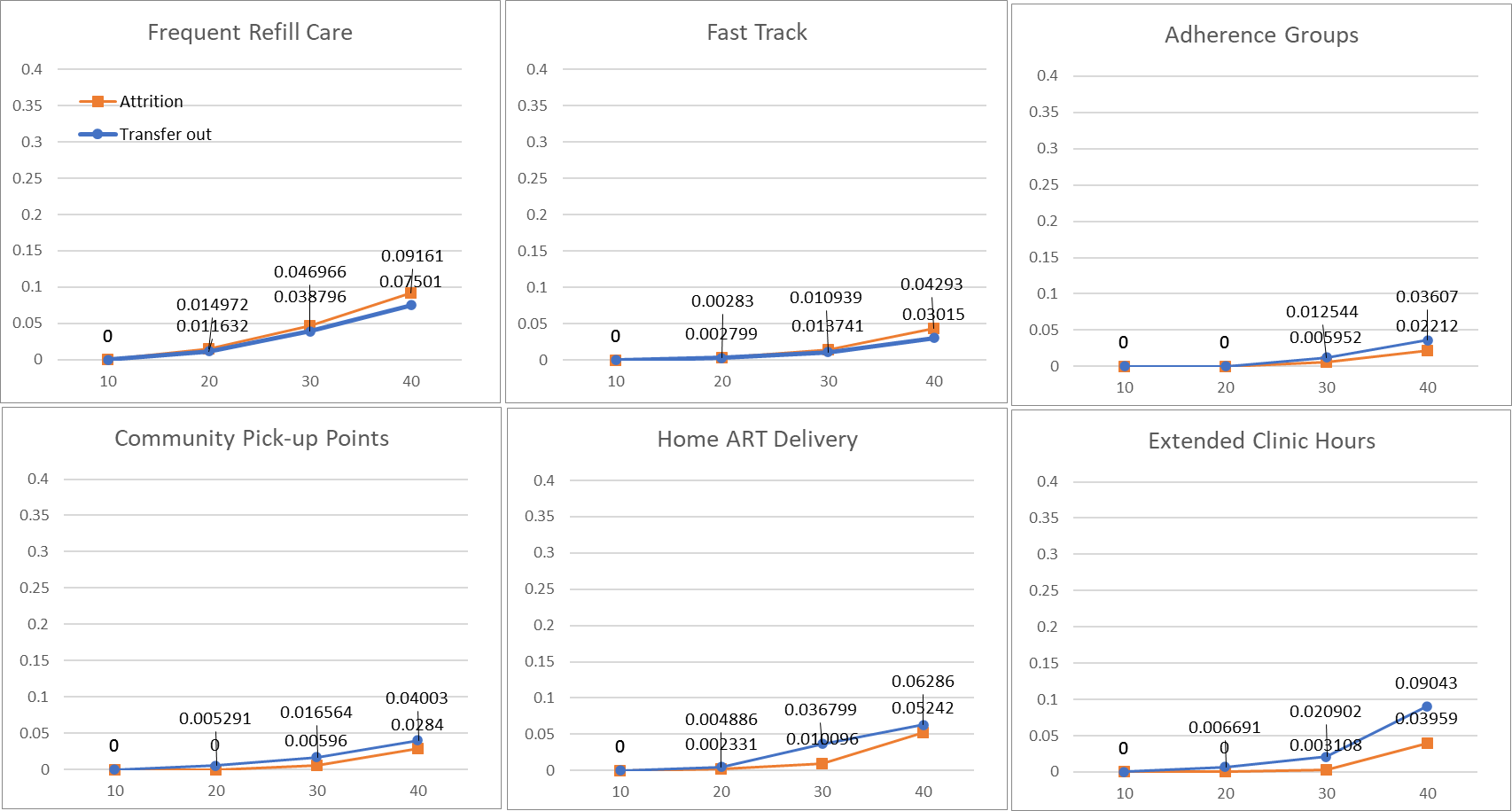

3) **4-6 months dispensing**

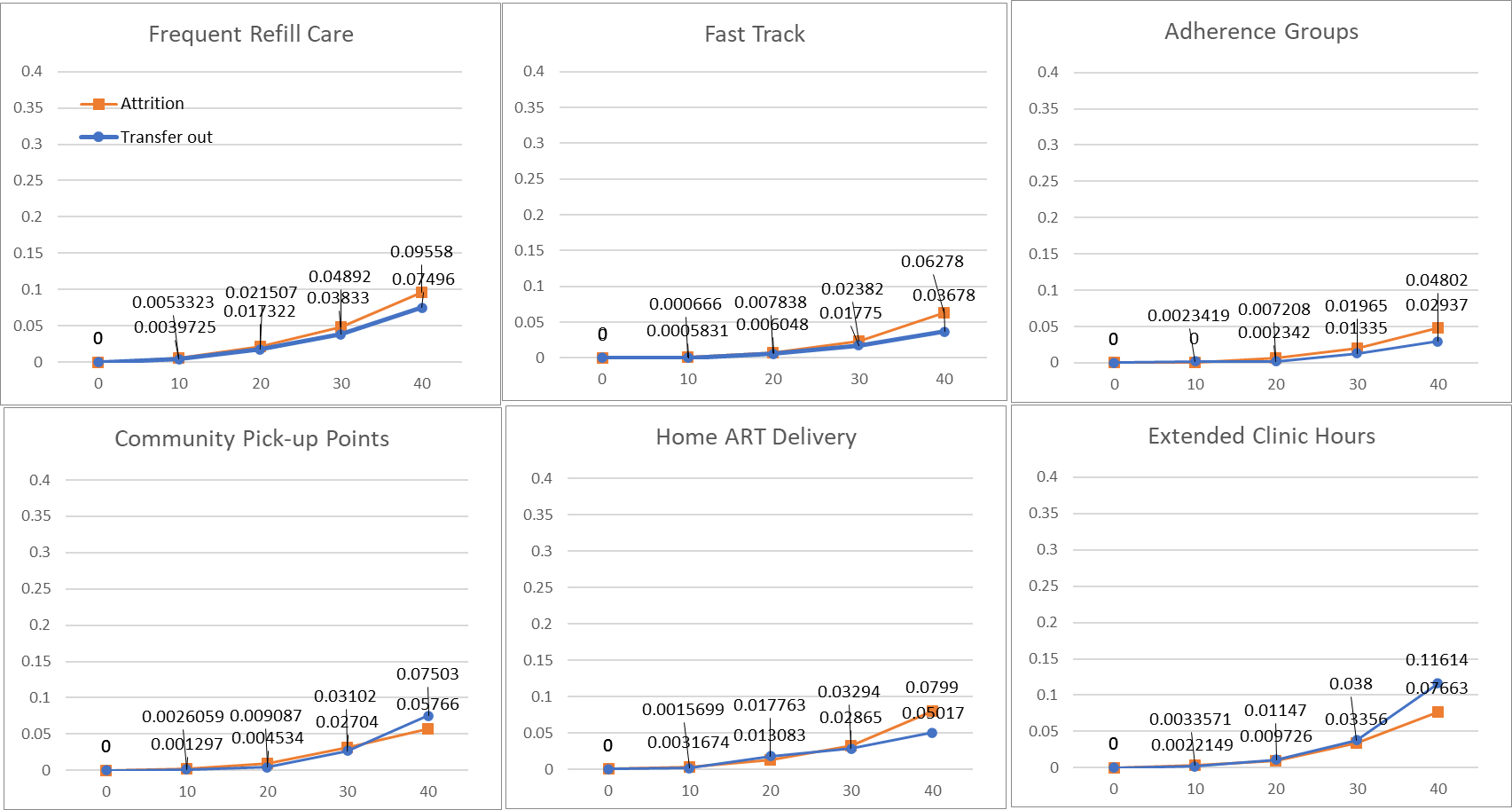

4) **Urban female**

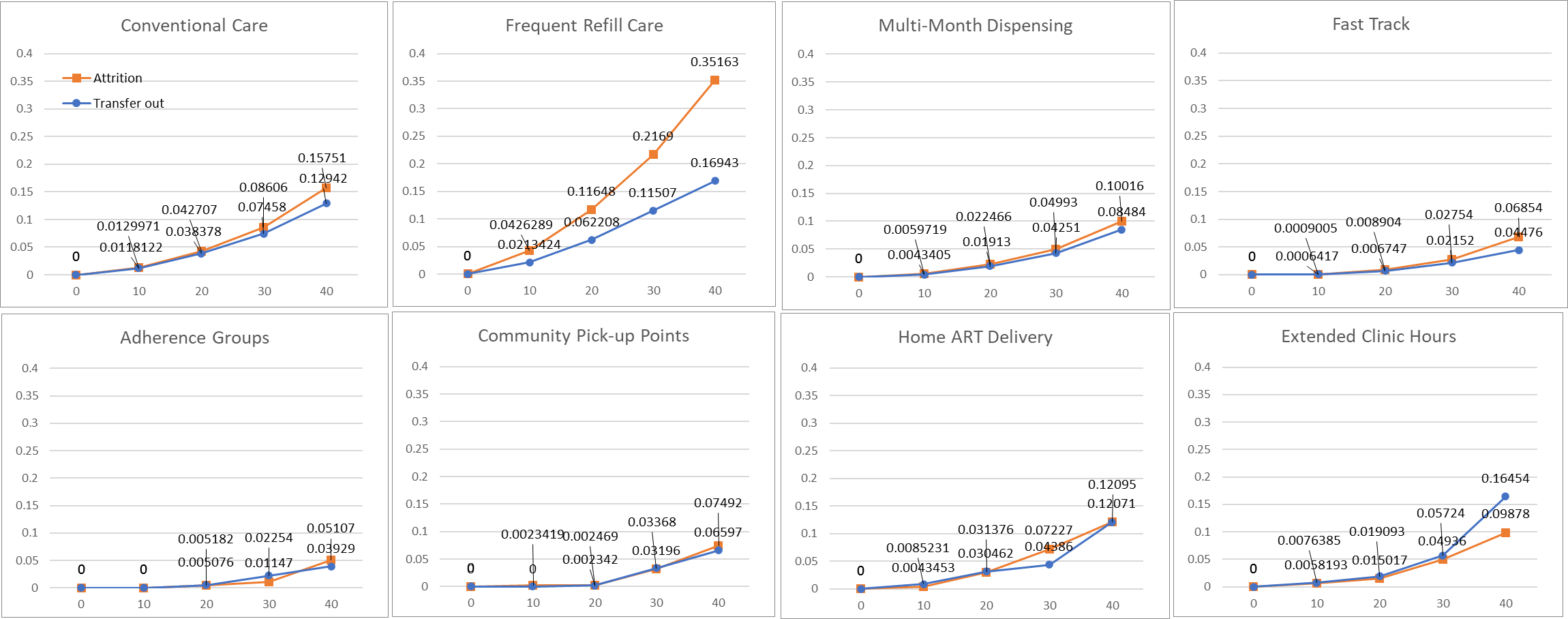

5) **Urban male**

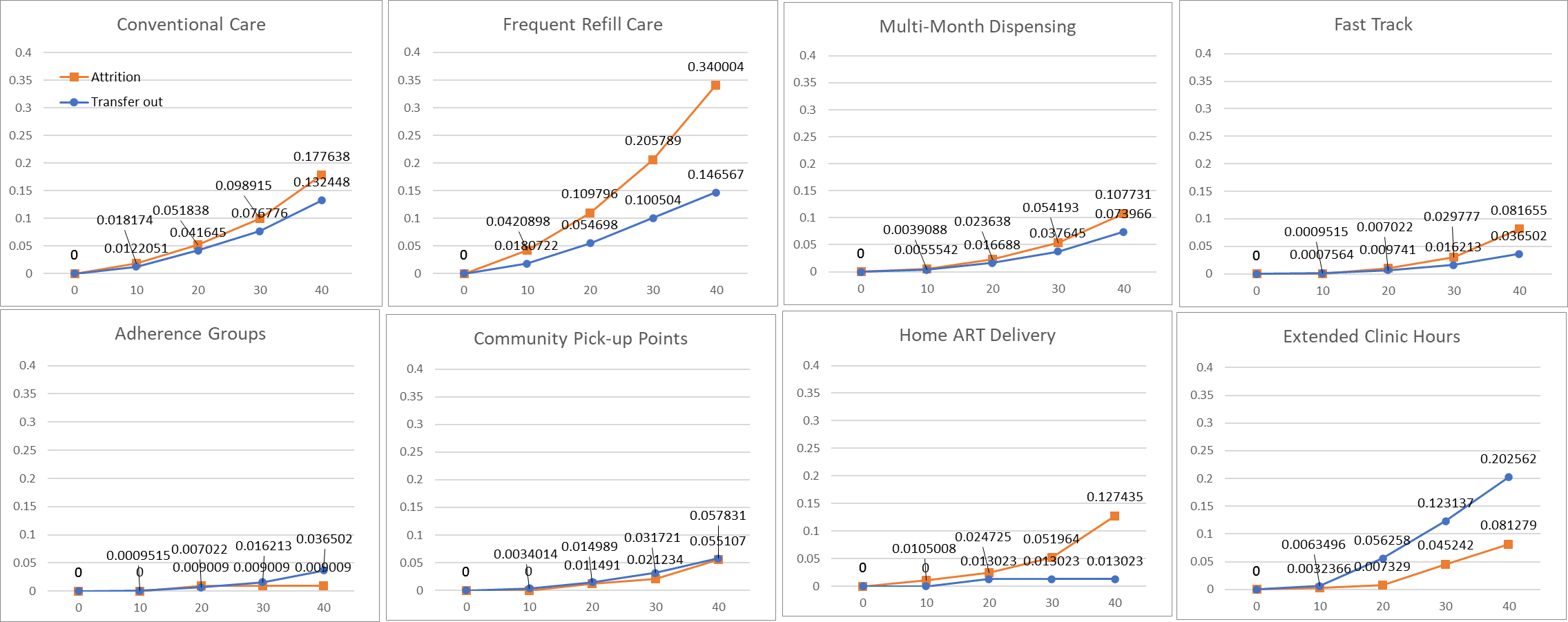

6) **Rural female**

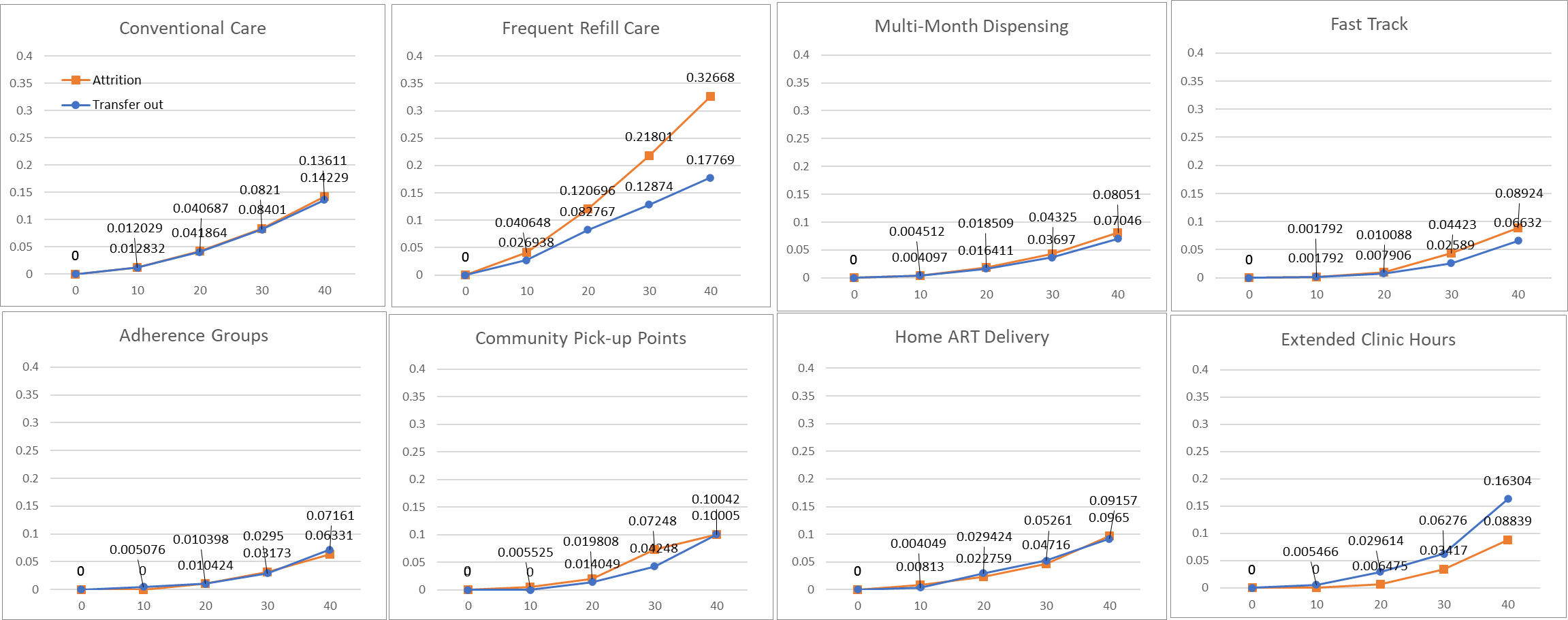

7) **Rural male**

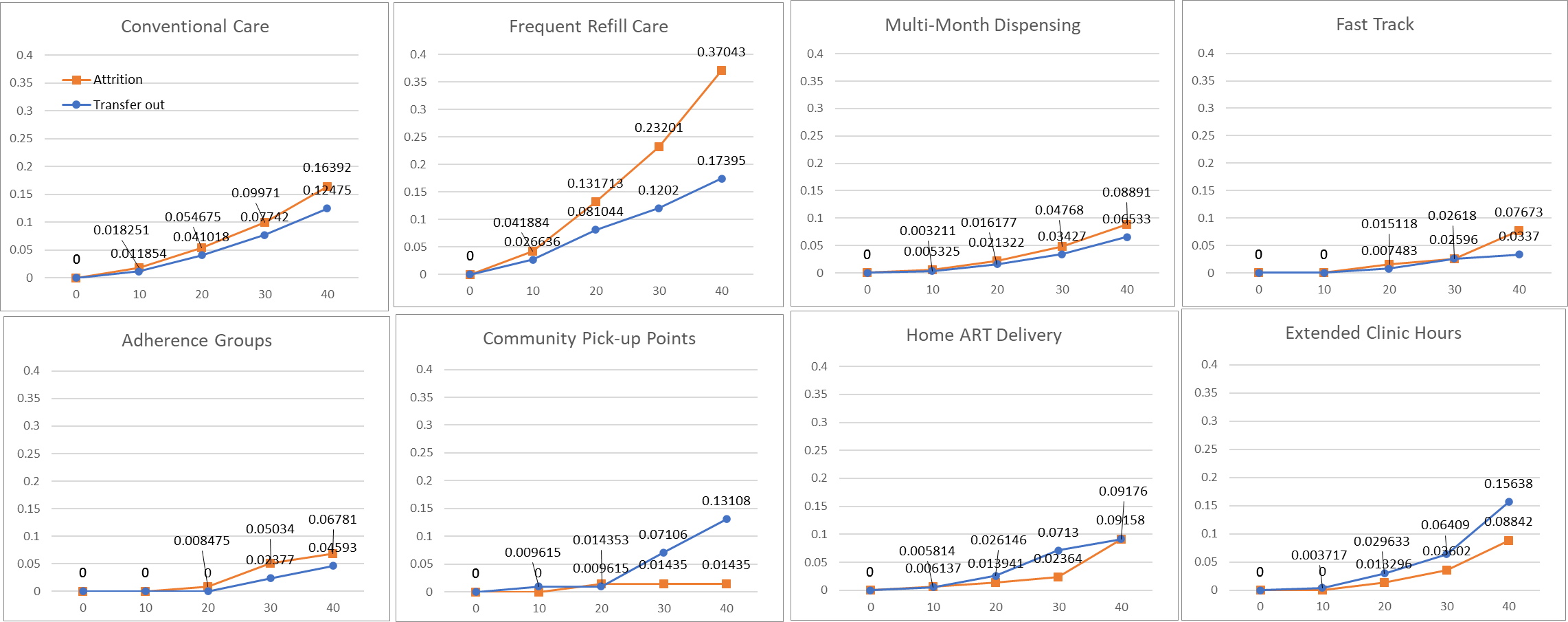
